# Preservation solutions modulate hydrogen sulfide synthesis in saphenous vein endothelium during coronary artery bypass grafting

**DOI:** 10.64898/2026.07.08.26357593

**Authors:** Matheus Duarte Pimentel, José Glauco Lobo Filho, Heraldo Guedis Lobo Filho, Emílio de Castro Miguel, Sergimar Kennedy de Paiva Pinheiro, Francisco Vagnaldo Fechine Jamacaru

**Affiliations:** Department of Surgery, Federal University of Ceará, Fortaleza, Brazil; Department of Metallurgical Engineering and Materials (DEMM) and Analytical Center, Federal University of Ceará, Fortaleza, Brazil; Nucleus of Research and Development of Medicines, Federal University of Ceará, Fortaleza, Brazil

**Keywords:** saphenous vein, coronary artery bypass grafting, preservation solutions, vascular endothelium, hydrogen sulfide

## Abstract

**Background:** The saphenous vein (SV) remains the most widely used graft in coronary artery bypass grafting (CABG). However, graft failure over the years has compromised long-term outcomes. Preservation of the vascular endothelium is fundamental for vein graft patency, and hydrogen sulfide (H_2_S) a protective gasotransmitter, plays a significant role in vascular homeostasis. This study evaluated how different intraoperative preservation solutions modulate H_2_S-synthesizing enzymes and endothelial integrity.

**Methods:** SV segments from 20 CABG patients were subdivided into five groups: Control (immediate fixation), normal saline (NS; 0.9% NaCl), autologous heparinized arterial blood (AHB), histidine-tryptophan-ketoglutarate (HTK) solution, and a damage group (no solution for 30 minutes). Structural integrity was evaluated by measuring endothelial coverage using light microscopy, and the expression of eNOS, CD31, and H_2_S pathway enzymes (CSE, CBS, and 3-MPST) was assessed by immunofluorescence (IF) and confocal microscopy to determine mean fluorescence intensity (MFI).

**Results:** LM analysis revealed that AHB (89.66% ± 3.02) and HTK (88.72% ± 3.07) preserved endothelial coverage significantly better than NS (78.06% ± 4.48) and the Damage Group (76.82% ± 4.90; p < 0.001). In IF, all interventions reduced eNOS and CD31 expression compared to the control, but AHB and HTK maintained significantly higher levels than NS (p < 0.001). All three H_2_S-producing enzymes were detected in the GSV endothelium, with CSE being the most expressed isoform. The use of NS caused a marked depletion of these enzymes, while AHB and HTK showed specific superiority in preserving H_2_S synthesizing enzymes.

**Conclusions:** The choice of preservation solution significantly affects endothelial integrity and the modulation of enzymatic H_2_S synthesis. NS proved to be deleterious to the endothelium, whereas AHB and HTK better preserved vascular structure and function, suggesting their clinical superiority for the preparation of venous grafts during CABG.

**Clinical Perspective:** *What Is New?:*

- This study confirms the presence of the three primary hydrogen sulfide (H_2_S)-synthesizing enzymes—cystathionine γ-lyase (CSE), cystathionine β-synthase (CBS), and 3-mercaptopyruvate sulfurtransferase (3-MPST)—in the human saphenous vein (SV) endothelium.
- It demonstrates that the use of normal saline (0.9% NaCl) as a storage solution for vein grafts—a common practice in many surgical centers—promotes a considerable reduction in the expression of H_2_S and nitric oxide (NO)-producing enzymes and negatively impacts the physical integrity of the endothelium.
- HTK solution and autologous blood preserve not only the endothelial structure but also the production of essential gasotransmitters, specifically and NO, within the vein graft.

*What Are the Clinical Implications?:*

- The proper choice of intraoperative preservation solution should be standardized to avoid the use of normal saline, aiming to minimize initial endothelial dysfunction.
- Maintaining H_2_S expression using buffered solutions or autologous blood may be a noteworthy strategy to preserve the SV endothelium during coronary artery bypass grafting (CABG).
- Preserving these gasotransmitter pathways can mitigate intimal hyperplasia and accelerated atherosclerosis, ultimately improving long-term vein graft patency and clinical outcomes for patients

## Introduction

Coronary artery bypass grafting (CABG) remains the treatment of choice for complex forms of coronary artery disease (CAD).^1,2^ The saphenous vein (SV) remains widely utilized in CABG, yet it exhibits limited long-term patency, primarily due to the development of vein graft disease (VGD). This pathophysiological process is characterized by intimal thickening and accelerated atherosclerosis, which can ultimately lead to vein graft failure (VGF). ^3–6^

Maintaining endothelial integrity starting from the intraoperative period is an important factor in reducing VGD rates; therefore, strategies to minimize intraoperative damage to the SV have become increasingly relevant. ^7–9^

During CABG, the SV graft is habitually subjected to a period of *ex-vivo* ischemia, during which it is stored in a preservation solution until the moment of anastomosis. The choice of this preservation solution has been a subject of debate for decades.^10–12^ Normal saline (NS; 0.9% NaCl) is frequently used, despite evidence suggesting that its acidic pH (∼5.0) and the absence of physiological components can be deleterious to the endothelium.^10,12,13^ Conversely, the use of autologous heparinized arterial blood (AHB) or balanced crystalloid solutions, such as histidine-tryptophan-ketoglutarate (HTK), has gained prominence due to their potential to mitigate oxidative stress and preserve vascular ultrastructure.^10,11,14–16^ In this context, selecting superior graft preservation solutions represents a simple, low-cost approach supported by experimental and clinical evidence of endothelial preservation.

Preservation solutions protect endothelial integrity through direct action on the cells of this vascular layer and by influencing the expression of vasodilatory, antithrombotic, and antioxidant substances.^5,10,13^ Notable among these are nitric oxide (NO), a well-established mediator of vascular homeostasis, and hydrogen sulfide (H_2_S). In recent years, H_2_S has been the subject of numerous studies illustrating its role in various physiological functions, and it is currently considered the third gasotransmitter alongside carbon monoxide (CO) and NO.^17–21^

Specifically in the vascular endothelium, physiological H_2_S expression acts in a complementary and synergistic manner with NO. It has the potential to prevent atherogenic changes in low-density lipoproteins (LDL), reduce monocyte adhesion, and promote vascular relaxation. Furthermore, H_2_S limits the development and progression of intimal hyperplasia by blocking the migration and proliferation of vascular smooth muscle cells. It also reduces vascular atherosclerosis and exerts antithrombotic and antiplatelet effects.^17,18,21,22^

Enzymatic H_2_S production in the vascular endothelium is predominantly mediated by three enzymes: cystathionine β-synthase (CBS), cystathionine γ-lyase (CSE), and 3-mercaptopyruvate sulfurtransferase (3-MPST).^18,21,23^ The synthesis of these enzymes can be affected by factors such as changes in pH and osmolarity, the bioavailability of substrates like homocysteine and cystathionine, and the presence of reactive oxygen species (ROS).^17,21,24^

Preservation solutions used in CABG may modulate the endothelial capacity for H_2_S and NO production, which would result in a direct effect on the patency of venous grafts used for myocardial revascularization.^10,20,25,26^ According to the current literature, this is the first study to evaluate the effect of different preservation solutions on the modulation of the H_2_S enzymatic pathway in the human SV endothelium.

The objective of this study was to evaluate the effect of different preservation solutions on the modulation of H_2_S synthesis in the SV endothelium during CABG. We specifically investigated how these solutions affect the expression of the enzymes CSE, CBS, and 3-MPST, aiming to identify the strategy that best preserves the expression of this gasotransmitter in the vein graft.

## Methods

### Ethical Aspects

This study was conducted in accordance with the Declaration of Helsinki and was approved by the Research Ethics Committee of the Federal University of Ceará under protocol numbers 4.978.835 and 7.351.596. All patients provided written informed consent prior to inclusion in the study.

### Study Design and Patient Selection

This is an *ex-vivo*, prospective, and controlled comparative study. We enrolled 20 patients, aged 45 to 80 years, who underwent elective CABG between February 2023 and April 2024. Inclusion criteria comprised patients of both sexes with an indication for elective CABG and no prior diagnosis of chronic venous disease. To ensure the quality of the conduits, all SV were evaluated by preoperative Doppler ultrasound and direct intraoperative visualization to confirm the absence of varicosities, reflux, or surgical trauma.

### Graft Harvesting and Surgical Protocol

The harvesting of the venous grafts was performed by a single surgeon using atraumatic techniques, including skin-bridged incisions and controlled intraluminal distension limited to a maximum of 100 mmHg. Surplus vein segments were harvested and immediately divided into subsegments of approximately 0.5 cm each. These segments were paired, ensuring that five segments from the same patient were distributed across the five experimental groups.

### Experimental Groups and Preservation Solutions

The 100 vein segments were allocated into five study groups (n=20 per group):

1. **Negative Control Group (Absent Damage):** Segments were immediately fixed upon surgical excision in a solution of 4% paraformaldehyde and 2.5% glutaraldehyde in 0.1 M sodium cacodylate buffer (pH 7.4), serving as the baseline reference for structural integrity.
2. **Normal Saline (NS) Group:** Samples were maintained in 0.9% sodium chloride for 30 minutes.
3. **Autologous Heparinized Arterial Blood (AHB) Group:** Segments were preserved for 30 minutes in the patient’s own arterial blood, collected with the addition of heparin to prevent clot formation.
4. **HTK Group:** Grafts were stored for 30 minutes in Custodiol® (histidine-tryptophan-ketoglutarate), a commercial organ preservation solution.
5. **Positive Control/Damage Group (Present Damage):** Segments were kept in room air for 30 minutes without any preservation solution to assess the impact of *ex-vivo* handling and tissue dehydration.

All preservation solutions were maintained at room temperature to mimic standard intraoperative conditions during CABG.

### Tissue Processing

Following the 30-minute experimental interval, all segments from the NS, AHB, HTK, and Damage groups were fixed in 4% paraformaldehyde and 2.5% glutaraldehyde in 0.1 M sodium cacodylate buffer (pH 7.4). Subsequently, specimens were dehydrated in increasing concentrations of ethanol and infiltrated with historesin (Leica, DE). Transverse sections of 5.0 μm were obtained using a rotary microtome (RM2265, Leica, DE) and mounted on glass slides for subsequent analysis.

### Light Microscopy and Endothelial Integrity

For light microscopy analysis, specimens were stained with 1% toluidine blue. This metachromatic dye was selected for its high affinity for acidic tissue components, such as nucleic acids and sulfates, allowing for precise visualization of nuclei and the endothelial cell body against the extracellular matrix. Qualitative and quantitative analysis was conducted by an examiner blinded to the experimental groups using a Primo Star light microscope (Zeiss, DE). Images were captured at 10x and 40x magnifications using ZEN Microscopy software (Zeiss, DE).

Endothelial integrity was quantified through digital morphometric analysis using ImageJ software (NIH, Bethesda, USA). The total perimeter of the venous lumen was outlined, and the length of the luminal surface covered by intact endothelial cells was subsequently calculated. The result was expressed as the percentage of total endothelial coverage (length of intact endothelium / total lumen perimeter x 100). Additionally, specific pathological changes were identified, such as cellular vacuolization, intimal layer detachment, and areas of endothelial denudation with eventual red blood cell adhesion.

### Immunofluorescence and Confocal Microscopy Tissue Preparation and Permeabilization

For in situ protein detection, 5.0 μm transverse sections mounted on glass slides were subjected to an indirect immunolabeling protocol. Initially, samples were rehydrated in phosphate-buffered saline (PBS, pH 7.4) for three 5-minute cycles. Cell membrane permeabilization was performed with 0.1% Triton X-100 (Merck, DE) for 15 minutes at room temperature, followed by additional PBS washes.

### Blocking of Nonspecific Binding and Antibody Protocol

To minimize background noise and prevent nonspecific immunoglobulin binding, sections were incubated with 2% bovine serum albumin (BSA) (ImunoScan, BR) diluted in PBS for 60 minutes. Subsequently, monoclonal primary antibodies were applied at a 1:200 dilution for the individualized detection of CBS (sc-133154, Santa Cruz Biotechnology, Inc., USA), CSE (sc-374249, Santa Cruz Biotechnology, Inc., USA), 3-MPST (sc-376168, Santa Cruz Biotechnology, Inc., USA), nitric oxide synthase 3 (NOS3/eNOS; sc-376751, Santa Cruz Biotechnology, Inc., USA), and CD31 (sc-13537, Santa Cruz Biotechnology, Inc., USA). Incubation occurred for 3 hours in a dark humid chamber at room temperature.

After rigorous PBS washes, sections were exposed to the secondary antibody Alexa Fluor 568 IgG (Goat anti-mouse) (A-11004, Invitrogen, Thermo Fisher Scientific, Rockford, IL) at a 1:200 dilution for 2 hours under light protection. Slides were mounted with Fluoromount-G antifade medium (Invitrogen, Thermo Fisher Scientific, USA) and sealed with glass coverslips to preserve the fluorescent signal.

### Confocal Microscopy and Image Acquisition Standardization

Analysis was conducted using a Zeiss LSM-710 confocal microscope (Zeiss, DE). To ensure quantitative comparability between experimental groups, acquisition parameters were standardized: a helium-neon laser (543 nm) was used for excitation, with images obtained using the same detector gain, digital offset, and digital gain settings for all analyses. High-resolution images (1024 x 1024 pixels) were captured at 10x and 20x magnifications, focusing specifically on the endothelial layer.

### Digital quantification and quality controls

The magnitude of protein expression was determined by Mean Fluorescence Intensity (MFI) using ImageJ software (NIH, Bethesda, USA). Regions of Interest (ROIs) were established and specifically delineated over the luminal endothelium. For each segment, the final MFI was calculated by subtracting the signal intensity obtained from negative controls (slides processed with the omission of the primary antibody, containing only the secondary antibody), thereby eliminating interference from tissue autofluorescence and nonspecific fluorophore binding. Quantitative analysis was performed in a blinded manner by an independent examiner to mitigate interpretation bias.

Detailed descriptions of the immunofluorescence protocols, antibody validation, confocal microscopy metadata, and expanded histomorphometric results are provided in the Supplemental Material available with this article.

### Statistical analysis

Continuous variables, including the percentage of endothelial coverage of the venous lumen and the MFI, were expressed as mean ± standard deviation (SD). The assumption of data distribution normality was formally tested and confirmed across all groups using the Kolmogorov-Smirnov test. For the comparison of means among the five experimental groups (Control, NS, AHB, HTK, and Damage), one-way analysis of variance (ANOVA) was utilized. In cases where ANOVA indicated significant global differences, Tukey’s post-hoc test was applied to perform multiple pairwise comparisons and identify specific differences between groups. All analyses were performed using two-tailed tests, and statistical significance was defined as a p-value < 0.05. Statistical procedures and the preparation of graphs were processed using GraphPad Prism software version 10.0 (GraphPad Software, San Diego, CA, USA) and Microsoft Excel (Microsoft Corporation, USA)

## Results

### Study patient characteristics

Twenty patients who underwent elective CABG between February 2023 and April 2024 were included in this study. The mean age of the cohort was 64.7 ± 10.13 years, with a male predominance (75%). The prevalence of cardiovascular comorbidities was considerable: 90% of patients had systemic arterial hypertension and 85% had a diagnosis of dyslipidemia. Half of the study population had triple-vessel coronary artery disease, and the mean EuroScore II was 0.96% ± 0.52. Detailed demographic and preoperative clinical data are described in Table 1.

**Table 1.** Demographic and preoperative clinical characteristics of the twenty patients studied.

| Variable | N |
| --- | --- |
| Age (years) (mean ± SD) | 64,7 ± 10,13 |
| Age > 75 years | 4 (20%) |
| Female sex | 5 (25%) |
| Male sex | 15 (75%) |
| Previous MI | 1 (5%) |
| CHF with Functional Class III/IV (NYHA) | 0 |
| BMI (kg/m<sup>2</sup>) (mean ± SD) | 27,59 ± 3,35 |
| Smoking history | 6 (30%) |
| Alcohol consumption history | 3 (15%) |
| COPD | 2 (10%) |
| Non-insulin-dependent diabetes | 8 (40%) |
| Insulin-dependent diabetes | 0 |
| Kidney failure | 0 |
| Dyslipidemia | 17 (85%) |
| Systemic Arterial Hypertension | 18 (90%) |
| Left main disease | 4 (20%) |
| Two-vessel disease | 10 (50%) |
| Three-vessel disease | 10 (50%) |
| EF ≤ 35% | 0 |
| Cerebrovascular disease | 1 (5%) |
| Carotid artery disease (lesion ≥ 70%) | 0 |
| EuroScore II (mean ± SD) | 0,96% ± 0,52 |

MI: myocardial infarction; CHF: congestive heart failure; NYHA: New York Heart Association; BMI: body mass index; COPD: chronic obstructive pulmonary disease; EF: ejection fraction; EuroScore: European System for Cardiac Operative Risk Evaluation

### Endothelial Integrity and Coverage Analysis

In the 100 venous segments studied, LM analysis revealed that the preservation strategy directly influences SV architecture. The Control Group exhibited continuous endothelium and preservation of the media and adventitia layers (coverage: 91.93% ± 2.57). In contrast, the use of NS and the absence of solution (Damage Group) resulted in extensive cytoplasmic vacuolization, intimal detachment, and significant endothelial denudation.

Quantitatively, the percentage of luminal endothelial coverage was significantly lower in the NS (78.06% ± 4.48) and Damage (76.82% ± 4.90) groups compared to the Control (*P* < 0.001). Strategies based on the use of AHB (89.66% ± 3.02) and HTK solution (88.72% ± 3.07) preserved the physical integrity of the endothelium superior to NS (*P* < 0.001), showing no statistically significant difference compared to the control group (*P* = 0.30 and *P* = 0.058, respectively). Additional data regarding light microscopy ananalysis is avai

### Endothelial functional and integrity markers

MFI, the parameter used to measure eNOS and CD31 markers, showed a statistically significant reduction (*P* < 0.001) in the study groups compared to the control group, with these changes being minimized in the AHB and HTK groups.

### eNOS expression

In the Control Group, eNOS showed homogeneous and continuous cytoplasmic staining along the entire endothelium, with an MFI of 53,180 ± 5,385. Exposing the grafts for 30 minutes revealed differences between preservation solutions; the use of AHB and HTK solution mitigated enzymatic loss, maintaining MFI levels of 43,323 ± 3,897 and 42,815 ± 4,349, respectively. No statistical difference was found between these two strategies (*P* = 0.99). NS promoted a marked reduction in the expression of this enzyme (MFI: 30,381 ± 4,213). This value was significantly lower than those of the AHB and HTK groups (*P* < 0.001). The most relevant reduction occurred in the Damage Group (MFI: 25,338 ± 3,359). Qualitatively, confocal microscopy revealed that NS induced a faint and fragmented fluorescence pattern, with extensive areas of protein expression interruption. These data are illustrated in Figure 1.

**Figure 1.**
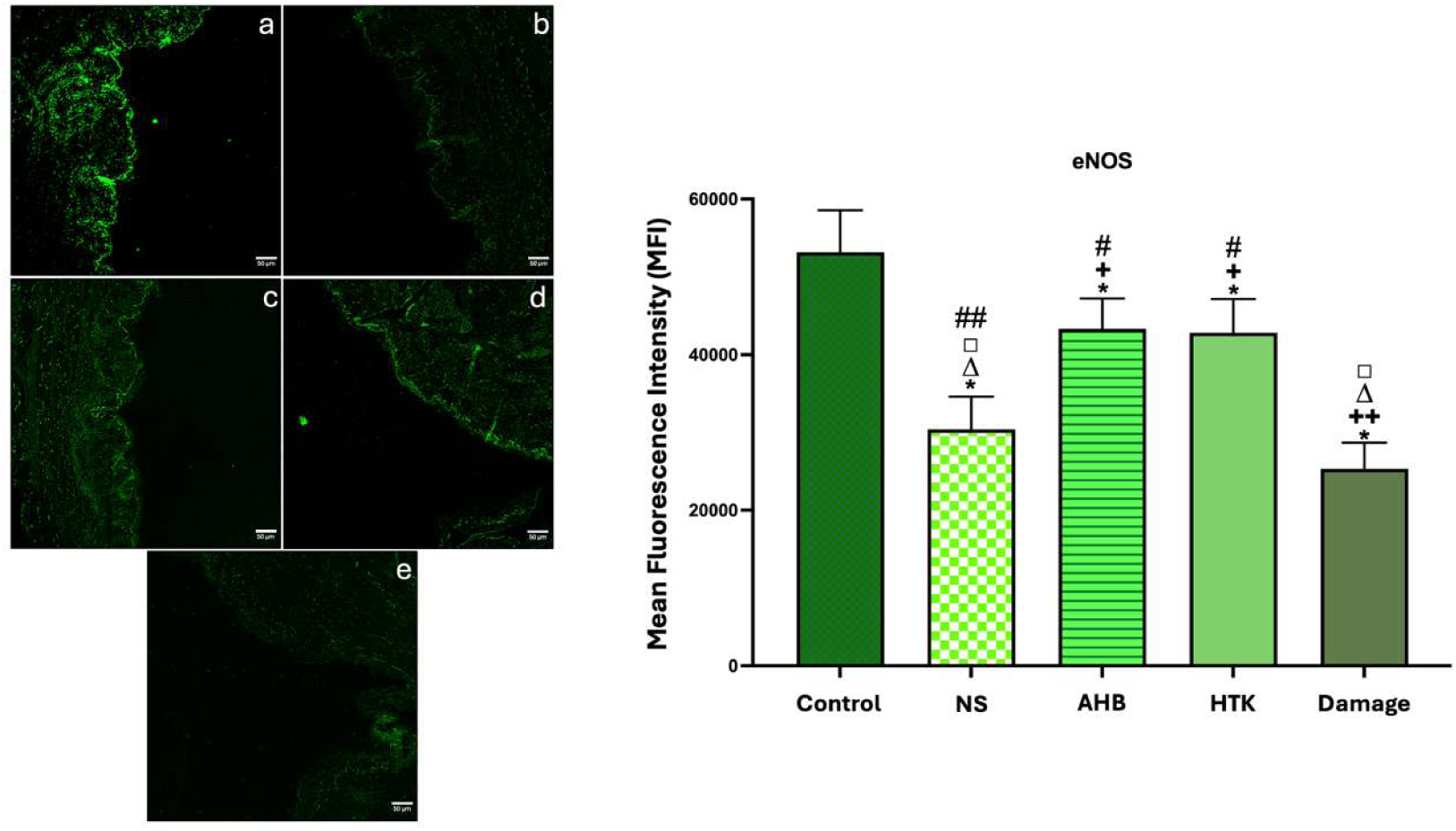
Effects of preservation solutions on eNOS expression in the saphenous vein (SV) endothelium. (A–E) Representative confocal microscopy images (10x magnification) showing immunostaining for eNOS (green) in the human saphenous vein (SV) endothelium. A: Control group; B: NS (Normal Saline) group; C: AHB (Autologous Heparinized Blood) group; D: HTK (histidine, tryptophan, and ketoglutarate) group; E: Damage group. To the right, a graph illustrating eNOS expression in the endothelium of twenty saphenous vein (SV) segments from each group. Data are expressed as mean ± standard deviation (SD) of the mean fluorescence intensity (MFI). The symbol * (*P* < 0.001) indicates statistically significant differences compared with the control group, whereas the symbols + (*P* < 0.001) and ++ (*P* = 0.003) indicate statistically significant differences compared with the NS group. The symbol Δ (*P* < 0.001) indicates statistically significant differences compared with the AHB group, the symbol □ (*P* < 0.001) indicates statistically significant differences compared with the HTK group, and the symbols # (*P* < 0.001) and ## (*P* = 0.003) indicate statistically significant differences compared with the Damage group. All pairwise comparisons between groups were performed using Tukey’s multiple comparison test.

### CD31 expression

CD31 staining in the Control Group also occurred homogeneously along the endothelial layer (MFI: 46,577 ± 5,159). The HTK solution presented an MFI of 38,646 ± 4,482, while AHB maintained 37,180 ± 3,259. Although the MFI in the HTK group was slightly higher, the difference between the two was not statistically significant (*P* = 0.77). Both groups demonstrated clear superiority in maintaining endothelial integrity when compared to NS. The use of NS resulted in an MFI of 26,275 ± 3,428, a value drastically lower than those of the balanced solutions (*P* < 0.001). The Damage Group recorded the lowest integrity (MFI: 22,306 ± 3,359). Qualitative analysis by confocal microscopy confirmed marked discontinuities in the luminal border of the NS and Damage groups, correlating with the areas of denudation observed by LM. These data are illustrated in Figure 2.

**Figure 2.**
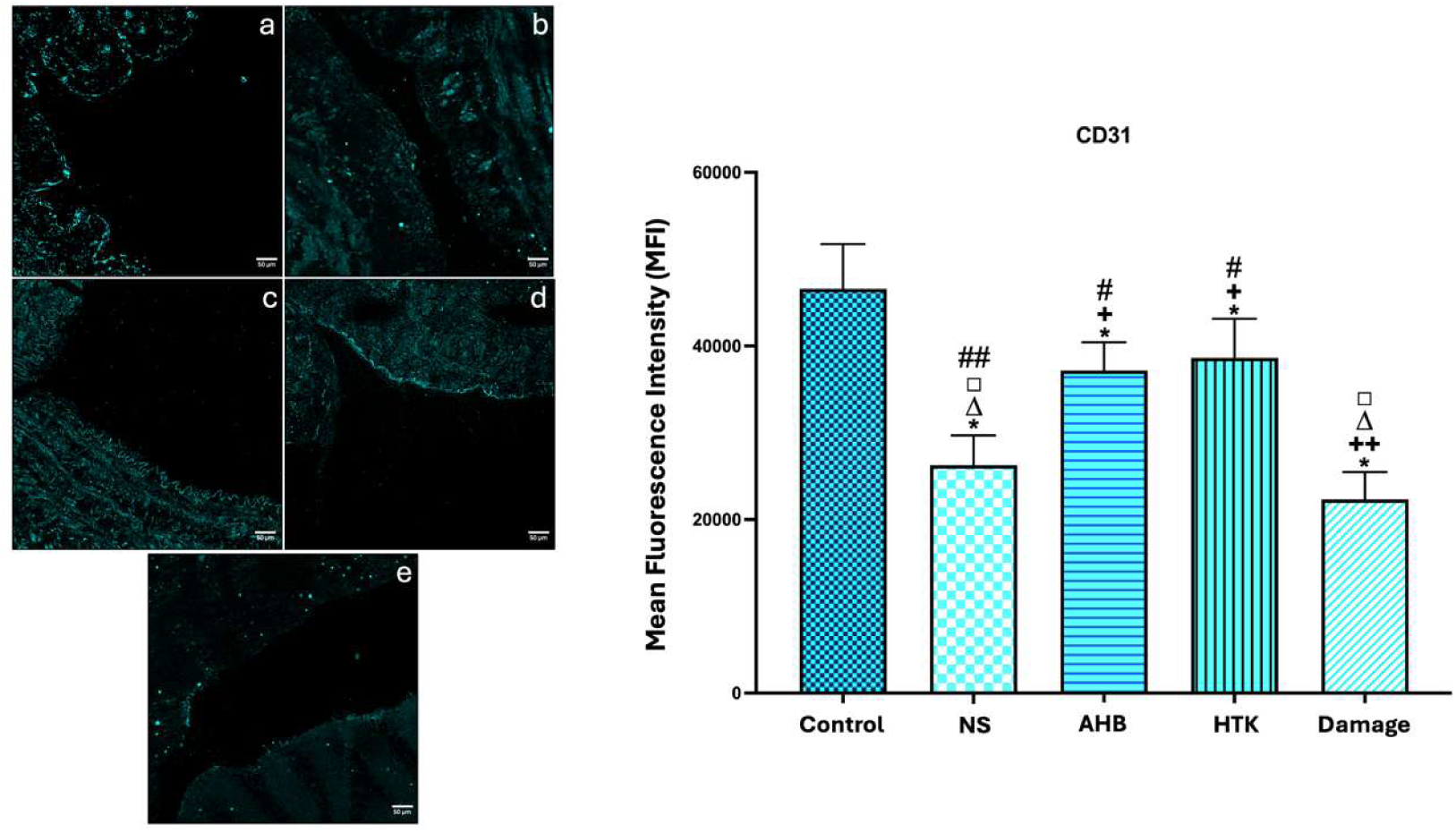
Effects of preservation solutions on CD31 expression in the saphenous vein (SV) endothelium. (A–E) Representative confocal microscopy images (10x magnification) showing immunostaining for CD31 (cyan blue) in the human saphenous vein (SV) endothelium. A: Control group; B: NS (Normal Saline) group; B: AHB (Autologous Heparinized Blood) group; D: HTK (histidine, tryptophan, and ketoglutarate) group; E: Damage group. To the right, a graph illustrating CD31 expression in the endothelium of twenty SV segments from each group. Data are expressed as mean ± standard deviation (SD) of the mean fluorescence intensity (MFI). The symbol * (*P* < 0.001) indicates statistically significant differences compared with the control group, while the symbols + (*P* < 0.001) and ++ (*P* = 0.017) indicate statistically significant differences compared with the NS group, the symbol Δ (*P* < 0.001) indicates statistically significant differences compared with the AHB group, the symbol □ (*P* < 0.001) indicates statistically significant differences compared with the HTK group, and the symbols # (*P* < 0.001) and ## (*P* = 0.017) indicate statistically significant differences compared with the Damage group. All pairwise comparisons between groups were performed using Tukey’s multiple comparison test.

### Modulation of enzymatic H_2_S expression

The presence of the three primary H_2_S-synthesizing enzymes was confirmed in the SV endothelium, with cystathionine γ-lyase (CSE) identified as the most expressed isoform under baseline conditions. There was a statistically significant reduction in mean fluorescence intensity (MFI) for all enzymes across all study groups compared to the control group (*P* < 0.001), with these changes being minimized in the AHB and HTK groups.

### Cystathionine γ-lyase (CSE)

In the Control Group, CSE exhibited intense and homogeneous endothelial staining (MFI: 37,163 ± 2,978). The use of AHB (30,137 ± 3,014) and HTK solution (31,457 ± 2,459) mitigated the reduction in enzymatic expression, maintaining MFI levels significantly higher than those observed with normal saline (NS) (20,840 ± 3,400) and in the Damage Group (17,267 ± 3,375) (*P* < 0.001 for both comparisons). No statistical difference was found between AHB and HTK in preserving CSE expression (*P* = 0.65). Qualitatively, the NS and Damage groups presented a fragmented and discontinuous fluorescence pattern, indicating depletion of the expression of this enzyme. These data are illustrated in Figure 3.

**Figure 3.**
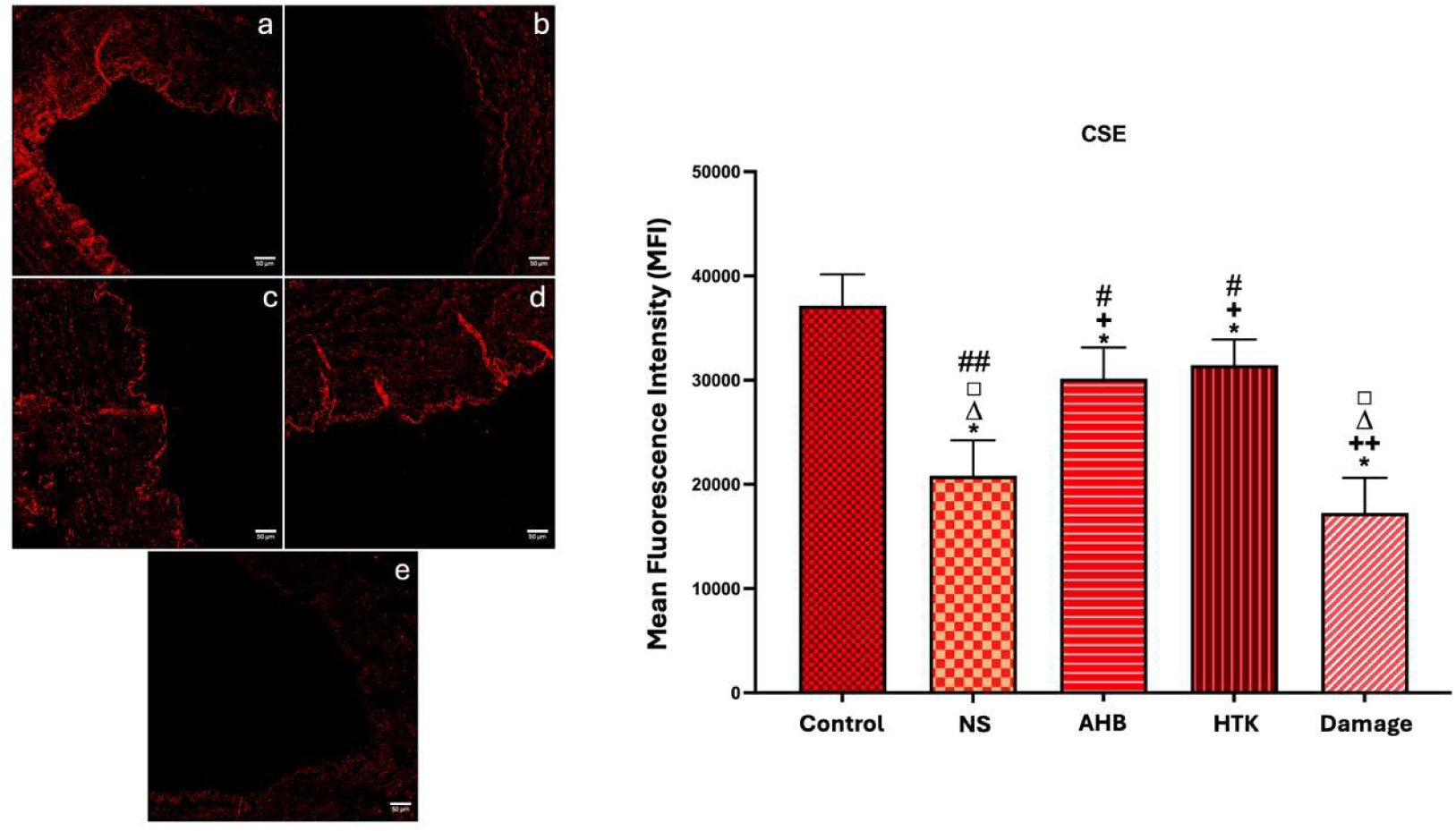
Effects of preservation solutions on cystathionine γ-lyase (CSE) expression in the saphenous vein (SV) endothelium. (A–E) Representative confocal microscopy images (10x magnification) showing immunostaining for CSE (red) in the human saphenous vein (SV) endothelium. A: Control group; B: NS (Normal Saline) group; C: AHB (Autologous Heparinized Blood) group; D: HTK (histidine, tryptophan, and ketoglutarate) group; E: Damage group. To the right, a graph illustrating CSE expression in the endothelium of twenty SV segments from each group. The symbol * (*P* < 0.001) indicates statistically significant differences compared with the control group, whereas the symbols + (*P* < 0.001) and ++ (*P* = 0.034) indicate statistically significant differences compared with the NS group. The symbol Δ (*P* < 0.001) indicates statistically significant differences compared with the AHB group, the symbol □ (*P* < 0.001) indicates statistically significant differences compared with the HTK group, and the symbols # (*P* < 0.001) and ## (*P* = 0.034) indicate statistically significant differences compared with the Damage group. All pairwise comparisons between groups were performed using Tukey’s multiple comparison test.

### Cystathionine β-synthase (CBS)

The MFI of CBS expression in the Control Group was 30,191 ± 3,602. Among the preservation strategies, HTK solution provided the greatest specific protection for this isoform, presenting an MFI of 27,229 ± 2,182, a value statistically superior to that observed in the AHB group (23,912 ± 2,343; *P* < 0.05). Both groups (HTK and AHB) were drastically superior to NS (15,775 ± 3,416) and the Damage Group (11,475 ± 1,964) (*P* < 0.001). Confocal analysis revealed that the HTK solution maintained the integrity of the fluorescent signal throughout the entire endothelial perimeter, while NS promoted areas of complete interruption of CBS expression. These data are illustrated in Figure 4.

**Figure 4.**
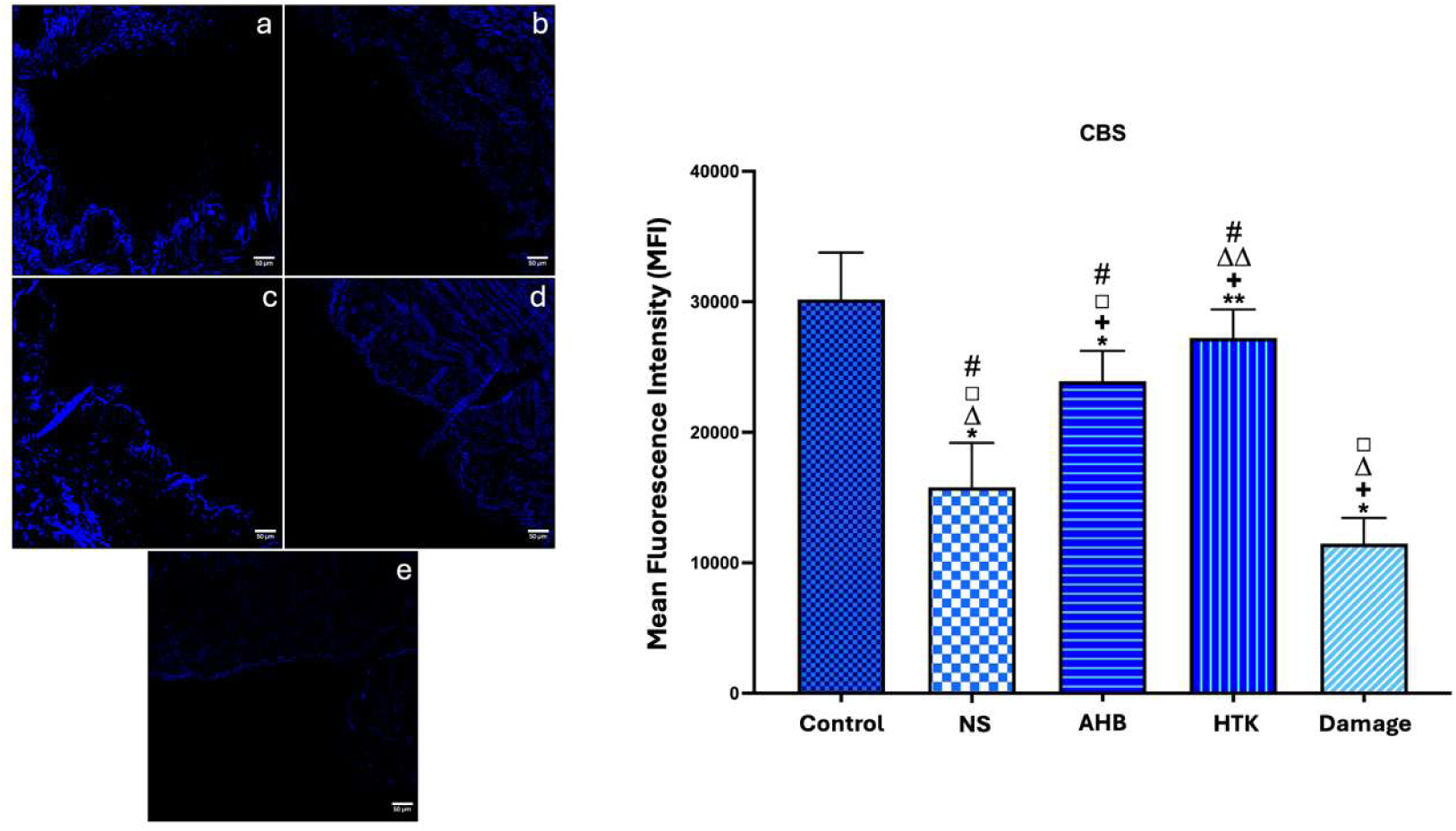
Effects of preservation solutions on cystathionine β-synthase (CBS) expression in the saphenous vein (SV) endothelium. (A–E) Representative confocal microscopy images (10x magnification) showing immunostaining for CBS (blue) in the human saphenous vein (SV) endothelium. A: Control group; B: NS (Normal Saline) group; C: AHB (Autologous Heparinized Blood) group; D: HTK (histidine, tryptophan, and ketoglutarate) group; E: Damage group. To the right, a graph illustrating CBS expression in the endothelium of twenty SV segments from each group. The symbols * (*P* < 0.001) and ** (*P* = 0.009) indicate statistically significant differences compared with the control group, while the symbol + (*P* < 0.001) indicates statistically significant differences compared with the NS group. The symbols Δ (*P* < 0.001) and ΔΔ (*P* < 0.0026) indicate statistically significant differences compared with the AHB group, the symbol □ (*P* < 0.001) indicates statistically significant differences compared with the HTK group, and the symbol # (*P* < 0.001) indicates statistically significant differences compared with the Damage group. All pairwise comparisons between groups were performed using Tukey’s multiple comparison test.

### 3-Mercaptopyruvate Sulfurtransferase (3-MPST)

For the 3-MPST enzyme, the Control Group presented an MFI of 33,946 ± 3,512. The pattern of enzymatic expression maintenance followed the trend of the previous isoforms: the HTK (28,813 ± 3,296) and AHB (26,378 ± 3,849) groups preserved enzymatic expression superiorly compared to NS (22,508 ± 2,888) and the Damage Group (17,459 ± 2,761) (*P* < 0.001). No significant difference was detected between HTK and AHB for this marker (*P* = 0.14). Staining in the control group and the balanced solutions (HTK/AHB) remained uniform, while segments preserved in NS exhibited a diffuse and low-intensity signal. These data are illustrated in Figure 5.

**Figure 5.**
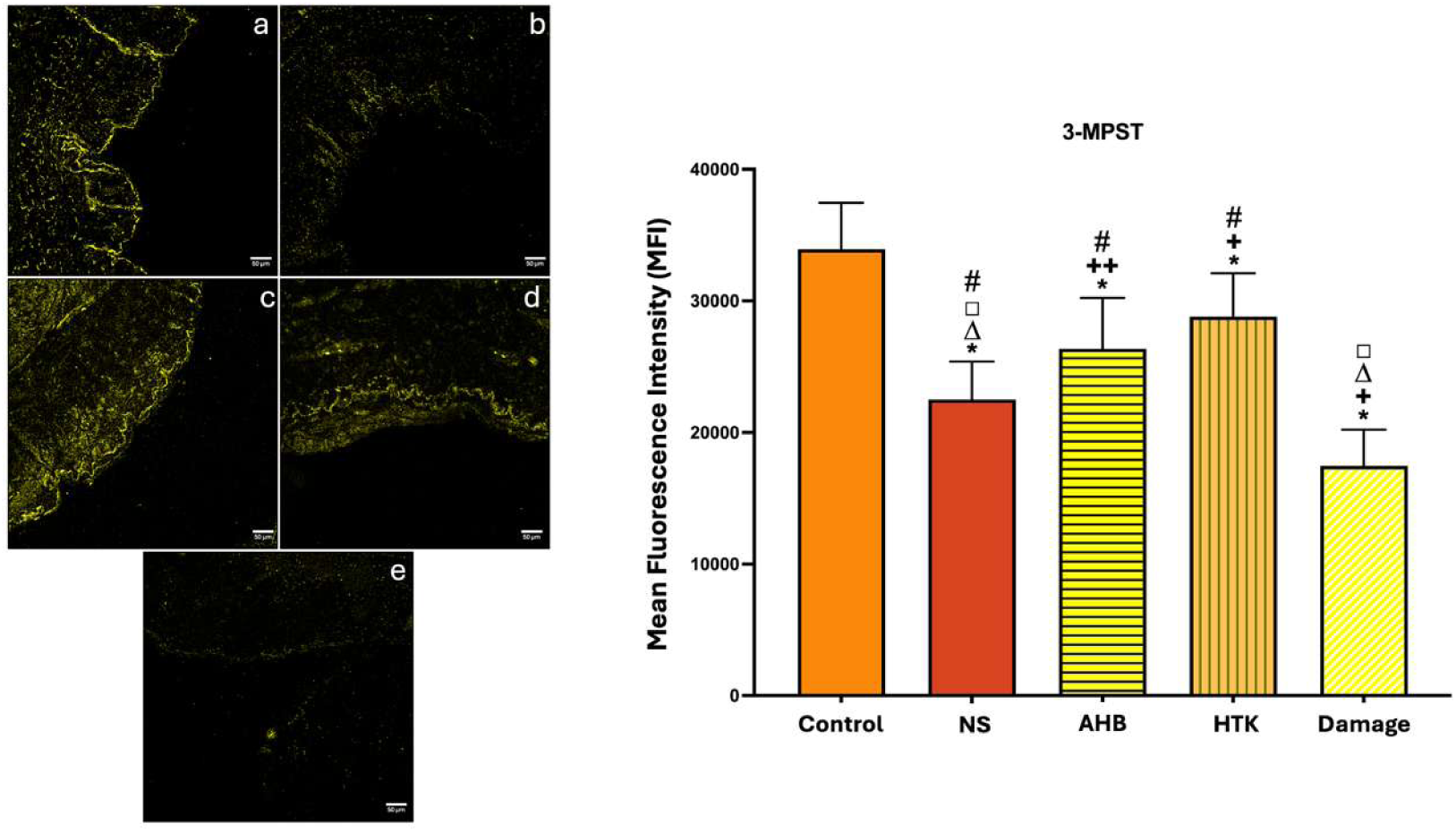
Effects of preservation solutions on 3-Mercaptopyruvate Sulfurtransferase (3-MPST) expression in the saphenous vein (SV) endothelium. (A–E) Representative confocal microscopy images (10x magnification) showing immunostaining for 3-MPST (yellow) in the human saphenous vein (SV) endothelium. A: Control group; B: NS (Normal Saline) group; C: AHB (Autologous Heparinized Blood) group; D: HTK (histidine, tryptophan, and ketoglutarate) group; E: Damage group. To the right, a graph illustrating 3-MPST expression in the endothelium of twenty SV segments from each group. The symbol * (*P* < 0.001) indicates statistically significant differences compared with the control group, whereas the symbols + (*P* < 0.001) and ++ (*P* = 0.003) indicate statistically significant differences compared with the NS group. The symbol Δ (*P* < 0.001) indicates statistically significant differences compared with the AHB group, the symbol □ (*P* < 0.001) indicates statistically significant differences compared with the HTK group, and the symbol # (*P* < 0.001) indicates statistically significant differences compared with the Damage group. All pairwise comparisons between groups were performed using Tukey’s multiple comparison test.

## DISCUSSION

The SV, after more than five decades of its standardization for use in CABG, remains an indispensable vascular graft.^3,4,8,9^ Continued attention throughout these years has been given to the preparation and intraoperative arrangements of venous grafts, aiming to provide adequate remodeling for their new function as an artery, especially through the search for atraumatic techniques that preserve endothelial integrity and the functionality of this tissue, maintaining the expression of vasodilatory, anti-inflammatory, and antithrombotic substances naturally expressed in this vascular layer, such as NO and H2S. ^8,10,13,14,17,27–30^

The relationship between H_2_S and the vascular endothelium has been increasingly explored in the literature, with descriptions of the protective effects of this gasotransmitter on vascular integrity and function.^17,21,27^ While there is evidence of H_2_S-producing enzyme expression in the internal thoracic artery and radial artery,^31^ in SV grafts, there is still a scarcity of evidence regarding how venous graft preparation methodologies during CABG can influence the expression of this molecule and the enzymes responsible for its synthesis.

The choice of the solution used for intraoperative graft preservation is, therefore, an aspect of considerable relevance for the maintenance of the vascular endothelium and a major focus of this study.^8,11,14,32^ This point has been discussed since the 1970s, with authors such as O’Connell and Gundry highlighting the effect of these solutions on endothelial structure and the early development of intimal hyperplasia.^12,33^

The effect of preservation solutions on endothelial cells is primarily due to their metabolic characteristics, since, unlike most cells in the human organism, those of the endothelium generate most of their ATP via the anaerobic pathway through glycolysis.^34,35^ This seemingly counterintuitive mechanism, being much less efficient than oxidative phosphorylation for energy production, proves to be a positive adaptive variation because the endothelium becomes less exposed to the production of ROS, more resistant to hypoxia, and better able to transfer O_2_ to the perivascular tissue cells.^34–37^

Endothelial tolerance to hypoxia is a crucial adaptive mechanism for the survival of cells in this layer, as reductions in arterial pressure are primarily sensed by this cell group. This resistance to depletion in endothelial cells is, however, dependent on the availability of glucose and other substrates for cellular metabolism, which are absent in preservation solutions such as NS.^14,34–37^

Consequently, although widely used, the 0.9% sodium chloride solution is associated with endothelial dysfunction due to its high osmolarity, acidity (pH around 5.0), and the absence of components that maintain endothelial cell viability while the vein graft is immersed in it.^10,11,38–40^

Preservation solutions for vascular grafts should ideally take these characteristics into account to be biocompatible with the endothelium. This has led to AHB and, more recently, balanced crystalloid solutions being chosen for the storage of vascular grafts, which also positively impacts synthesis.

Despite this evidence, it is worth noting that surgeons sometimes still do not consider the choice of preservation solution a relevant topic in CABG, which results in a lack of standardization in this aspect.^15,32,41^

In our study, we observed that the use of NS presented deleterious effects on endothelial integrity, with a considerable attenuation of the percentage of the vascular lumen covered by endothelium, in addition to marked reductions in the MFI levels of CD31 and eNOS. These aspects were statistically different from what was noted with the use of AHB and HTK, which showed a greater protective effect on the endothelium. This is in agreement with what is illustrated in the literature and with a recent study from our research group, which demonstrated that, regardless of the pressure used to distend the vein grafts, there are deleterious effects of NS use on the endothelial ultrastructure.^42^

Regarding synthesis, the results of this study demonstrate the expression of CSE, CBS, and 3-MPST in the SV endothelium. It was observed that the use of preservation solutions reduced the levels of these enzymes in the venous endothelium, with a considerably more pronounced depletion in the NS and Damage group segments. This suggests that in these groups, where there is greater oxidative stress, a higher degree of hypoxia, and more evident tissue damage, enzymatic expression became more compromised. This implies a consequent reduction in expression and increased endothelial vulnerability to the inflammatory and oxidative damage resulting from surgical excision and hypoxia, as well as a greater likelihood of subsequent endothelial dysfunction and VGF.

The use of AHB, in comparison to NS, demonstrated a protective effect on enzymatic expression, limiting the reduction in MFI levels, likely by reducing—at least during the short period the vein graft remained preserved in it—the degree of hypoxia and oxidative stress. This resulted in superior MFI levels of CSE, CBS, and 3-MPST compared to the NS and Damage groups.

The HTK solution presented favorable results in preserving both endothelial integrity and functionality. Regarding synthesis, it showed superiority in the detected levels of CSE, CBS, and 3-MPST compared to the NS and Damage groups. Additionally, it promoted better preservation of CBS levels relative to the SV group preserved in AHB, a finding potentially attributable to the previously described role of HTK in protecting cells from oxidative stress.^43^

## Limitations

Despite the methodological rigor employed, clinical studies, especially those conducted at a single center, generally allow for limitations in their findings. Quantification by MFI via confocal microscopy provides a robust estimate of *in situ* enzymatic expression but does not directly measure the dynamic production or the absolute tissue concentration of gasotransmitters, which would require methods such as gas chromatography. Additionally, the protocol of this study evaluated the damage resulting from 30 minutes of *ex-vivo* ischemic preservation; however, the clinical impact of these changes on graft patency was not analyzed.

Finally, we emphasize the pioneering nature of this investigation in characterizing the modulation of the enzymatic pathway in the human SV endothelium specifically within the context of CABG. Given the scarcity of previous evidence regarding the behavior of this gasotransmitter in venous grafts, additional limitations and biological variables not yet mapped may become evident as subsequent studies expand this line of research.

## Conclusion

The AHB and HTK solutions used in CABG for SV preservation positively modulate the expression of the three enzymes (CSE, CBS, and 3-MPST) responsible for synthesis in the venous endothelium. NS acts in a deleterious manner, compromising the structural integrity of the vein graft and reducing the enzymatic synthesis of H_2_S.

## AUTHORS CONTRIBUTIONS

**Matheus Duarte Pimentel:** Conceptualization; Data curation; Formal Analysis; Investigation; Methodology; Project administration; Resources; Visualization; Writing – original draft; Writing – review & editing. **José Glauco Lobo Filho:** Conceptualization; Data curation; Formal Analysis; Methodology; Supervision; Validation; Visualization; Writing – review & editing. **Heraldo Guedis Lobo Filho:** Conceptualization; Data curation; Formal Analysis; Methodology; Supervision; Validation; Visualization; Writing – review & editing. **Emílio de Castro Miguel:** Conceptualization; Data curation; Formal Analysis; Investigation; Methodology; Supervision; Validation; Visualization; Writing – review & editing. **Sergimar Kennedy Pinheiro Paiva:** Data curation; Formal Analysis; Investigation; Methodology; Visualization; Writing – review & editing.

## Data Availability

Data from this manuscript is available within the article.

## Nonstandard Abbreviations and Acronyms

3-MPST: 3-mercaptopyruvate sulfurtransferase
AHB: Autologous heparinized arterial blood
CABG: Coronary artery bypass grafting
CBS: Cystathionine β-synthase γ-lyase
CSE: Cystathionine γ-lyase
HTK: Histidine-tryptophan-ketoglutarate
MFI: Mean fluorescence intensity
NS: Normal saline
SV: Saphenous vein
VGD: Vein graft disease
VGF: Vein graft failure

